# Clinical and Demographic Characteristics of COVID-19 Patients Admitted in a Tertiary Care Hospital in the Dominican Republic

**DOI:** 10.1101/2020.12.11.20247437

**Authors:** David De Luna, Yori Roque, Nicolás Batlle, Katherine Gómez, Miguelina Jáquez, Brinia Cabrera, Lissa de la Cruz, Osvaldo Tavárez, Rossy Belliard, José Javier Sanchez

**Affiliations:** HOMS - COVID Study Team, Hospital Metropolitano de Santiago, Santiago de los Caballeros, Dominican Republic; Pontificia Universidad Católica Madre y Maestra, Santiago de los Caballeros, Dominican Republic

## Abstract

To present clinical and demographic characteristics of COVID-19 patients admitted to Hospital Metropolitano de Santiago in Dominican Republic, we analyzed electronic medical records of all hospitalized patients clinically admitted as viral pneumonia through March - April, 2020. Of 374 patients, 150 (40.1%) laboratory confirmed, were included in this study. Most of the patients were men (104 / 69.3%) with a median (IQR 44 - 66) age of 54. Hypertension (83 / 55.3%) and diabetes mellitus (49 / 32.7%) were the most common comorbidities, whereas fever (120 / 80%), cough (79 / 52.7%) and fatigue (60 / 40%) were the most common presenting symptoms. 28 (18.7%) patients required admission to the intensive care unit, of them, 26 patients (17.3%) required mechanical ventilation. The overall mortality rate was 10.7% Higher levels of inflammatory markers were associated with longer length of stay (LOS). This findings indulge information that could contribute to stratify patients at higher risk of complications.

## Background

By the end of 2019, the world began to confront numerous cases of viral pneumonia, initially detected in Wuhan, China, which quickly spread all around the globe, becoming a pandemic.^1^ Since its naming and further description, much is known about SARS-CoV-2 and its associated disease, COVID - 19, allowing health personnel a better understanding of its dynamics and outcomes. To date, almost a year after these first cases, more than 70 million people have been affected, and more thank 1,5 million have died.^2^ Recently, several drugs have demonstrated beneficial effects in COVID-19, including antivirals that can reduce viral replication and lessens the length of stay, but none have been able to prevent new infections or act as post exposure prophylactic agent.

The Dominican Republic (DR) has been struggling with COVID-19 since the first confirmed case back in March 2020, in an italian tourist in the northern part of the country. By then, the country was in the middle of an electoral process, which is believed contributed to the rapid propagation and community transmission of COVID-19. Even though several actions were taken, including a partial lockdown from dusk until dawn, the positivity rate rose considerably as soon as accurate diagnostic tests were more accessible to the population. Hospitals have been responding per demand, using different strategies to separate respiratory patients from non-respiratory. At the time this paper was submitted, near 150,000 cases have been diagnosed +, and more than 2,000 have died from the disease.^3^

In DR several third-level public hospitals were chosen as primary COVID-19 health care centers; however, private hospitals represent a greater percentage of COVID-19 beds since the beginning, and the rapid growth in the number of cases demanded more beds to be able every week through the course of the pandemic. The majority of cases have been reported in three of the most populated cities of the DR: Santo Domingo, which is the capital, Santiago de los Caballeros, the largest city in the Cibao region, and San Francisco de Macorís, head county of Duarte Province.^3^ Here, in this document, we present the demographics and clinical characteristics of the first COVID-19 patients admitted in an academic, third-level hospital in Santiago de Los Caballeros, along with an analysis of the relationship between laboratory data and medication used in each patient and their outcome.

## Methods

### Study design and participants

In this retrospective single-center study, all hospitalized patients clinically diagnosed with viral pneumonia through March - April at the Hospital Metropolitano de Santiago (HOMS), were initially included in this cohort. Hospital Metropolitano de Santiago is a third level academic hospital in Santiago De Los Caballeros and has been one of the main hospitals in the city and region designated and prepared to hospitalize COVID-19 patients. Pharyngeal swabs of these patients were used for SARS-CoV-2 detections. Those patients with negative results were excluded. The study was approved by the institutional review board (IRB) of Pontificia Universidad Católica Madre y Maestra (PUCMM).

Epidemiological, demographics, clinical (comorbidities, signs and symptoms), laboratory, management, and outcome data were obtained from patients’ electronic medical records and checked by two physicians. The date of the onset of COVID were recorded as well, defined as the first day when symptoms were noticed by the patients. Records, where there were missing data and clarification could not be made with the attending doctor or the health care providers were excluded.

### Laboratory procedures

Pharyngeal swabs were collected for the SARS-CoV-2 RNA detection through reverse transcriptase-polymerase chain reaction (RT-PCR). These reactions were performed in Laboratorio Nacional Dr. Defilló and Laboratorio Referencia, which were two of the three designated laboratories by the Public Health ministry in turn. Laboratory results on admission were collected for each patient, including hemoglobin, white blood cells count, platelets, renal and hepatic function tests, C-reactive protein (CRP), ferritin, lactate, dehydrogenase (LDH), D-dimer (DD) and serum creatine kinase (CK), as well as procalcitonin.

### Statistical analyses

Categorical variables were presented as frequencies and percentages, while continuous variables were summarized using median and interquartile range (IQR) values. Mann-Whitney test and chi-square were used to compare the variables for data from different patients groups with a 95% confidence interval (CI), as it corresponded. Pearson correlation coefficient was used to measure linear correlation between continuous variables. The data was analyzed in SPSS version 25.0.

## Results

Among 374 patients admitted as suspected cases of viral pneumonia, 150 (40.1%) cases confirmed through RT-PCR as COVID-19 were included in this study. Most of the patients were men (104 / 69.3%) and the median (IQR 44 - 66) age was 54. 67 (44.7%) patients reported comorbidities including hypertension (83 / 55.3%), diabetes mellitus (49 / 32.7%) and cardiovascular disease (13 / 8.7%). The median time from onset of symptoms to date of isolation was 6 days (IQR 3 - 9). Most of the patients (120 / 80%) reported fever as a presenting symptom, followed by cough (79 / 52.7%) and fatigue (60 / 40%). Other symptoms reported were myalgia (55 / 36.7%), dyspnea (50 / 33.3%), arthralgia (43 / 28.7%) and headache (42 / 28%). Diarrhea was reported by only 15 (10%) patients. During hospitalization, 28 (18.7%) patients were treated in the intensive care unit (ICU), of which, 26 patients (17.3%) required mechanical ventilation. The overall length of stay (LoS) was 7 days (IQR 5 - 10) reaching 13 days (IQR 8 - 38) in patients admitted to the ICU.

Of the 150 patients included in this cohort, 134 (89.3%) patients were discharged alive. The overall mortality rate was 10.7%. The median age of the patients who died was 64 years (IQR 58 - 75). These patients had higher levels of CRP, ferritin and LDH. Baseline laboratory findings are in table 2. Ferritin was found > 300 in a great proportion of patients, however, not other inflammatory marker was consistently out of range, but CRP in patients admitted to ICU.

**Table 1.** Demographic and baseline characteristics of patients with COVID-19.

| Characteristics | All patients (n=150) | Admission area |  |
| --- | --- | --- | --- |
|  |  | Respiratory ward (n=122) | Respiratory ICU (n=28) |
| **Sex — No. (%)** |  |  |  |
| Female | 46 (30.7) | 39 (31.9) | 7 (25) |
| Male | 104 (69.3) | 83 (68.0) | 21 (75) |
| **Age** |  |  |  |
| Median (IQR*) | 54 (44-66) | 54 (43-64) | 63 (46-72) |
| **Distribution — No. (%)** |  |  |  |
| 18 - 24 y | 3 (2) | 3 (2.4) | 0 (0) |
| 25 - 44 y | 39 (26) | 33 (27.0) | 6 (21.4) |
| 44 - 64 y | 66 (44) | 57 (46.7) | 9 (32.1) |
| ≥ 65 y | 42 (28) | 29 (23.7) | 13 (46.4) |
| **Comorbidity — No. (%)** | 67 (44.7) | 39 (68.0) | 10 (31.9) |
| Hypertension | 83 (55.3) | 68 (55.7) | 15 (46.4) |
| Diabetes Mellitus | 49 (32.7) | 43 (35.2) | 6 (21.4) |
| Heart disease | 13 (8.7) | 11 (9.0) | 2 (7.14) |
| Pulmonary disease | 6 (4) | 3 (2.4) | 3 (10.7) |
| Cancer | 5 (3.3) | 4 (3.2) | 1 (3.6) |
| Neurologic disease | 5 (3.3) | 4 (3.2) | 1 (3.6) |
| Reumathologic disease | 3 (2) | 2 (1.6) | 1 (3.6) |
| **Onset of symptoms prior admission - median (IQR\*)** | 6 (3-9) | 5.5 (3-14) | 6 (3-9) |
| **Symptoms on admission — No. (%)** |  |  |  |
| Fever | 120 (80) | 100 (81.9) | 20 (71.4) |
| Dyspnea | 50 (33.3) | 38 (31.1) | 12 (42.8) |
| Cough | 79 (52.7) | 67 (45.0) | 12 (42.8) |

\*IQR: interquartile range

| Characteristics | All patients (n=150) | Admission area |  |
| --- | --- | --- | --- |
|  |  | Respiratory ward (n=122) | Respiratory ICU (n=28) |
| Headache | 42 (28) | 34 (27.8) | 8 (28.5) |
| Myalgias | 55 (36.7) | 49 (40.2) | 6 (21.4) |
| Arthralgias | 43 (28.7) | 38 (31.1) | 5 (17.8) |
| Fatigue | 60 (40) | 52 (42.6) | 8 (28.5) |
| Diarrhea | 15 (10) | 12 (9.8) | 3 (10.7) |
| Others | 36 (24) | 27 (22.1) | 9 (32.1) |
| **Need of mechanical ventilation — No. (%)** | 26 (17.3) | 13 (10.6) | 13 (46.4) |
| **Length of Stay - Median (IQR\*)** | 7 (5 - 10) | 6 (4 - 9) | 13 (8 - 38) |
| **Death - No. (%)** | 16 (10.6) | 6 (4.9) | 10 (35.7) |

*\*IQR: interquartile range*

**Table 2.** Lab tests profile on admission

|  | Normal Range | All patients (n=150) | Admission area |  |
| --- | --- | --- | --- | --- |
|  |  |  | Respiratory ward (n=122) | Respiratory ICU (n=28) |
| **Median (IQR)\*** |  |  |  |  |
| Hemoglobin | 12-17 g/dL | 13.4 (12.3 - 14.4) | 13.4 (12.5 - 14.3) | 13.0 (11.5 - 14.1) |
| WBC | 5.00 - 10.00 cel/ml | 6.2 (4.5 - 8.6) | 6.0 (4.5 - 7.9) | 7.7 (4.6 - 11.5) |
| Platlets | 150.00 - 450.00 cel/dL | 224 (191-283) | 226 (190 - 280) | 214 (194 - 292) |
| Alanine aminotranferase | 13.00 - 39.00 U/L | 31.9 (20.3 - 53.9) | 30.0 (19.9 - 53.9) | 35.5 (22.1 - 86) |
| Aspartate aminotranferase | 7 - 52 U/L | 37.9 (26.7 - 54.1) | 36.4 (25.9 - 50.4) | 45.8 (34.4 - 64.3) |
| Blood urea nitrogen | 7.00 - 25.00 mg/dL | 13 (9.0-19.0) | 12.0 (9.0 - 17.0) | 19.0 (11 - 26) |
| Creatinine | 0.60 - 1.20 mg/dL | 0.9 (0.8 - 1.2) | 0.9 (0.8-1.2) | 0.9 (0.7 - 1.4) |
| C-Reactive Protein | 0.00 - 6.00 mg/L | 4.1 (1.7 - 8.7) | 3.5 (1.3 - 7.3) | 7.2 (4.1-14.1) |
| Ferritin | 11.00 - 306.80 ng/mL | 369 (170 - 670) | 358 (146 - 592) | 508 (215 - 1000) |
| Lactate | 0.90 - 1.70 mmol /L | 2.4 (1.9 - 3.4) | 2.4 (2.1 - 3.1) | 2.9 (1.9 - 4.1) |
| Total Creatin Kinase | 39.0 - 308 U/L | 120 (75 - 193) | 120.9 (78.7 - 180) | 120 (68 - 289) |
| Procalcitonin | 0.00 - 0.9 ng/mL | 0.12 (0.05 - 0.43) | 0.10 (0.05 - 0.30) | 0.22 (0.05 - 0.81) |
| D-Dimer | 0.00 - 0.50 µ/ml | 0.45 (0.26 - 0.80) | 0.77 (0.24 - 0.65) | 0.77 (0.32 - 3.76) |
| Lactate Dehydrogenase | 140 - 271 U/L | 274 (209 - 378) | 261 (193 - 350) | 315 (379 - 559) |

\*IQR: interquartile range

**Table 3.**
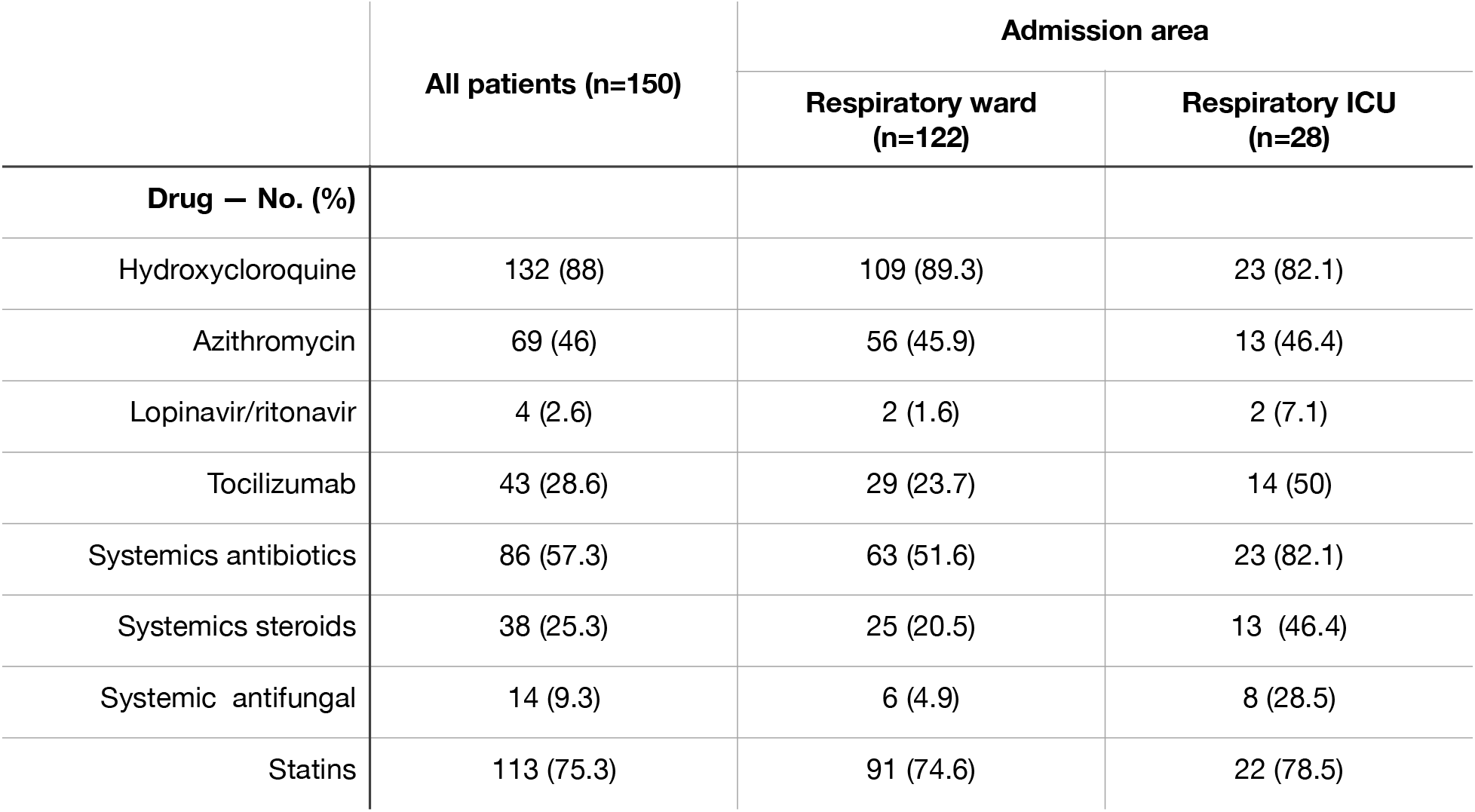
Distribution of different medications used to treat COVID-19 patients

**Table 4.**
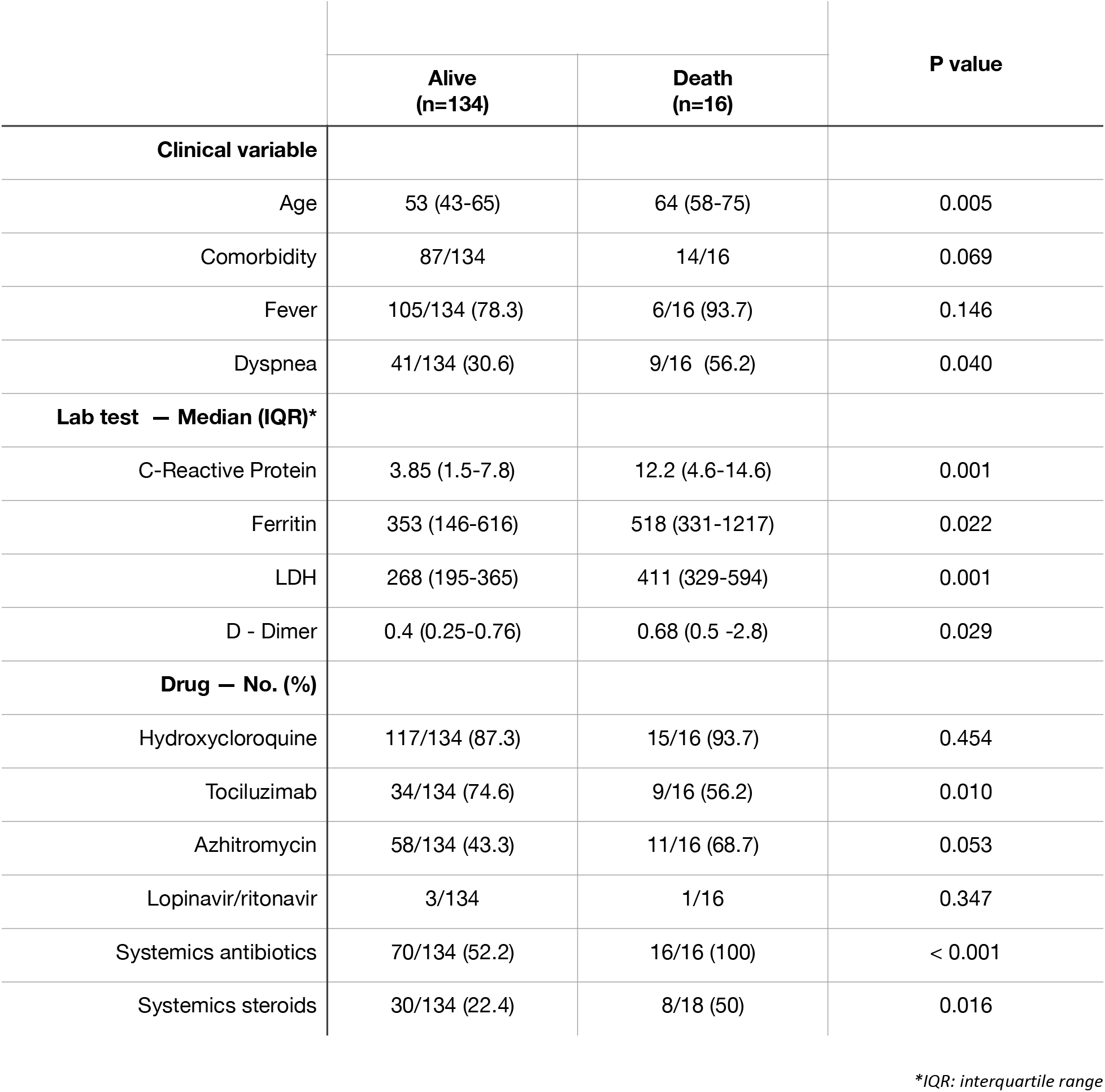
Correlation between clinical characteristics and discharge status

Among virus-targeted medication, most of the patients received Hydroxychloroquine (88%). Lopinavir/ritonavir was used in just 4 (2.6%) patients. The IL-6 inhibitor, tocilizumab, used as a host-targeted medication, was administrated in 43 (28.6%) patients, 14 were in ICU, while systemics steroids were used in 38 (25.3%) patients. Azithromycin was given to 69 (46%). Other antibiotics were administered in 86 patients (57.3%), 63 (51.6%) out of 122 were in the respiratory ward, while 23 (82.1%) out of 28 patients admitted in ICU received antibiotics as well. Antifungals were used in 14 (9.3%) patients, 8 (57.1%) were in ICU. Most of the patients (75.3%), received statins as part of the standard of care.

## Discussion

Back in March, RT-PCR was not widely available in the DR, which in turn delayed early diagnosis and isolation of positive COVID-19 cases. Many patients were admitted as viral pneumonia based on clinical suspicion and as they reported closed contact with diagnosed patients. Among all patients, 40.1% could be confirmed through RT-PCR. In line with several other reports, the majority were men (4,5,6). The median age of all patients was 54. Taking in consideration the median age in the DR is 28 years old^4^, this suggests older people are more at risk of being hospitalized as they get COVID. This data is very close to the results by Zhang et al. (57y) ^5^, but older than the population included in the report by Guang et al (47y)^6^, both in China, and younger than the report in New York by Richardson et al. (63y)^7^. The median age in both countries is around 38 years old^4^, which also goes in line with a higher risk for older people.

In this cohort, 67% of the patients have at least 1 comorbidity, in line with data reported by Grasselli et al. (68%)^8^. Hypertension was the most common comorbidity, similar to other reports, ^7, 9, 10^ followed by diabetes which was the same as data reported in NY and China.^7,10^ No obesity or immunodeficiency was observed in this cohort, which have been linked to complications in other reports.^11^

The median time for onset of symptoms was larger than what was reported by Chen *et al*.^12^ (6 days vs 4 days), with fever as the most common presenting symptom, in both groups of patients admitted in the respiratory ward and those who required ICU, in line with reports in Italy and various countries across the globe.^13,14^ Other symptoms commonly reported in patients requiring ICU were cough and dyspnea, while asthenia and myalgias were more reported in the patients who were not transferred to ICU. Unlike what was reported by Lechien et al.^15^, headache was reported much less than their European population, while their other main symptoms (loss of smell and taste dysfunction) were not reported in our cohort.

The proportion of patients who required ICU and mechanical ventilation was higher in this series than in other reports^6^. As expected, these patients had a longer LoS and increased risk of fatality outcome as they were prone to need more aggressive interventions, beyond antivirals and supportive care. Overall mortality was 10.7% quite similar to that reported by Goyal et al. (10.2%) in NY^11^. The median age for those who died was 64 years old. This agrees with data from Brazil ^16^, where a cross-sectional observational study showed a mean age of non-survivors of 65.3 years old, and survivors tended to be younger and were less likely to have any comorbidity. In our cohort, besides comorbidity, factors associated with fatality were elevated levels of CRP, ferritin, LDH and DD. Also, those patients who needed tocilizumab, antibiotics and systemic steroids as treatment, were more likely to die in ICU.

An interesting finding in this study was there were not remarkable alterations in laboratory counts among all patients. Leukopenia and thrombocytopenia were the most common alterations reported in many other series ^9, 11, 12^, and regarding inflammatory markers, elevations in CRP, ferritin a DD, have been associated with severe COVID and a higher risk of ventilation or ICU admission. Indeed, we found those who were admitted to ICU, tended to have higher ferritin and CRP (median 508 and 7.2, respectively) than those who did not require ICU (median 358 and 3.5, respectively), but the values were not as high as other reports.^7,12^

As the pandemic arouse, the urgency to have an effective medication to treat this virus, brought hydroxychloroquine to the front line, after it was rapidly identified as a potential drug because of its action against Middle East Respiratory Syndrome and severe acute respiratory syndrome *in vitro*. Based on the results by Gautret et al ^17^, it was used as an in-hospital medication, according to FDA’s recommendations after it was granted its’ emergency used authorization. Almost all patients included in this study received hydroxychloroquine as a virus-targeted medication, alone or together with azithromycin, while a small group received lopinavir/ritonavir, as there was some data^18^ regarding its’ potential activity against SARS-CoV-2 and its’ inclusion in the national protocol for diagnosis and treatment of COVID by the Public Health Ministry. None QT prolongation or other arrhythmias were seen associated with these drugs.

Regarding host-targeted medication, the IL-6 inhibitor tocilizumab was used in this population, as did systemic steroids as well. This was before Roches’ announcement^19^ about the outcomes of COVACTA study; by then, China approved its use, and some hospitals reported its’ efficacy in treating severe COVID.^20,21^ Systemics steroids were selected in patients with previous lung diseases and those with acute respiratory distress associated with pulmonary fibrosis due to COVID-19.

An important number of patients received antibiotics and antifungals, mostly as bacterial coinfection was suspected. According to Lai et al. ^22^, bacterial co-infection is very variable among COVID-19, but it can reach over 50% among non-survivors; *Streptococcus pneumoniae, Staphylococcus aureus, Klebsiella pneumoniae, Mycoplasma pneumoniae, Chlamydia pneumonia, Legionella pneumophila* and *Acinetobacter baumannii* were reported as the most common pathogens isolated from different hospitals in China, the US and Italy. Additionally, *Candida species, Aspergillus flavus* and other respiratory virus. In another report, Lansbury et al.^23^ found that only 7% of hospitalized patients had bacterial co-infection, increasing to 14% in ICU patients. Even though the frequency could be low, selection of adequate combination therapy to treat both bacteria and viruses represents a challenge, mainly as inflammatory markers would be elevated because of COVID-19, and the lacking procalcitonin usual efficacy to make a differential diagnosis of co-infection and superinfection between bacteria and other pathogens in this scenario, particularly because of the blocking effect of interferon-gamma and IL-1beta of its synthesis, as normal immune response occurs ^24^; Statins have been used in COVID-19 patients, according to evidence that suggests its potential role as adjunctive therapy to mitigate dysregulated inflammation and endothelial dysfunction ^25^.

There are two significantly important findings regarding distribution by discharge status; patients who were discharged alive were less likely to received tocilizumab, systemic steroids or antibiotics. This proportion of patients also tend to be younger. We hypothesize deaths associated with administration of any of these drugs, represent a group of patients with a poor baseline prognosis, and may be associated with a hyperinflammatory response induced by SARS-CoV-2.

## Conclusion

Clinical investigation of this group of patients provides information that can help clinicians to identify and stratify those patients at higher risk of complications, also analyzing the strategies applied at the beginning of this pandemic to develop better approaches to obtain best results.

## Data Availability

N/A

## Acknowledgments

The team would like to thank Marcos Tavarez, PhD, for his support reviewing this manuscript.

